# Association Between Out-of-Hospital Falls and Cardiovascular Events in Patients with Coronary Heart Disease: A Retrospective Cohort Study

**DOI:** 10.64898/2026.07.24.26358896

**Authors:** Changjiang Deng, Yuhang Men, Xinshu Xu, Yujie Pang, Zihan Tang, Haowen Ren, Weiyan Cui, Junrui Hou, Maimaitiming Muyesaier, Zhihang Chen, Haoyu Chen, Ting-Ting Wu

## Abstract

**Background:** Falls are increasingly recognized as adverse events in coronary heart disease (CHD) patients, yet their prognostic implications remain incompletely understood. This study examined the independent associations of falls wi th mortality and major adverse cardiovascular events, and the roles of functional status and frailty in these relationships.

**Methods:** This retrospective cohort study enrolled 2,139 CHD patients with median follow-up of 36 months. Falls were ascertained via telephone interviews every 3 months. Primary outcomes included major adverse cardiovascular eve nts (MACE) and major adverse cardiovascular and cerebrovascular events (MACCE). Secondary outcomes comprised all-cause and cardiac mortality. Multivariable Cox models with stepwise adjustment for functional status indicators were constructed, with subgroup analyses stratified by frailty status.

**Results:** During follow-up, 171 patients (8.0%) experienced falls. Falls were associated with mortality in univariate analysis but not after adjusting for functional status, indicating mediation by functional decline. In contrast, falls remained independently associated with MACE (HR=1.73, 95%CI: 1.17–2.57, P=0.0 06) and MACCE (HR=1.67, 95%CI: 1.14–2.46, P=0.009) in fully adjusted models. Frailty significantly modified this association (P for interaction <0.001). Among robust patients, falls conferred substantially elevated risk (MACE: HR =4.08, 95%CI: 2.37–7.01; MACCE: HR=3.94, 95%CI: 2.30–6.76), whereas no significant association was observed in pre-frail or frail patients.

**Conclusions:** Falls independently predict long-term MACE and MACCE in C HD patients, with mortality effects mediated by functional status. Frailty significantly modifies the fall-cardiovascular event relationship—robust patients experiencing falls face substantially elevated cardiovascular risk and warrant com prehensive evaluation. These findings support integrating fall history into cardiovascular risk assessment and implementing frailty-stratified management.

## Introduction

Coronary heart disease (CHD) remains a leading cause of mortality and morbidity worldwide(1–2). With population aging and improvements in acute-phase management, comprehensive long-term risk assessment has become increasingly important for CHD patients(3).

Falls represent a common but underappreciated adverse event in cardiovascular populations. Some research reports indicate that among patients who have recently been hospitalized due to cardiovascular diseases, over 50% have a significantly higher risk of falling, and this risk is independently correlated with a n increase in mortality rates and re-hospitalization rates(4–6). However, most previous studies have focused on fall risk assessment rather than actual fall events, and the relationship between falls and long-term cardiovascular outcomes in CHD patients remains incompletely characterized.

Patients with CHD have multiple predisposing factors for falls. Coronary artery disease itself has been identified as an independent predictor of falls, and comorbidities such as orthostatic hypotension may further impair balance function. Moreover, frailty—a syndrome of decreased physiological reserve—affects approximately 30% of older CHD patients and is closely associated with both fall risk and adverse cardiovascular outcomes. These observations suggest tha t frailty may play an important role in the relationship between falls and pro gnosis(7).

Despite these associations, several key questions remain unanswered. First, whether falls independently predict major adverse cardiovascular events (MACE) and major adverse cardiovascular and cerebrovascular events (MACCE) in CH D patients after accounting for traditional risk factors is unclear. Second, the pathways through which falls influence different endpoints—mortality versus cardiovascular events—have not been systematically evaluated. Third, whether frailty status modifies the association between falls and cardiovascular outcomes has not been adequately explored.

Therefore, this study aimed to: (1) determine the association between fall hist ory and long-term outcomes (all-cause mortality, cardiac mortality, MACE, an d MACCE) in CHD patients; (2) investigate the mediating role of functional status in these associations through stepwise adjustment models; and (3) explore effect modification by frailty status and other clinical characteristics through subgroup analyses.

## Methods

### Study design and population

This single-center retrospective cohort study enrolled patients aged ≥18 years with confirmed coronary heart disease (CHD) at the First Affiliated Hospital of Xinjiang Medical University between January 2018 and December 2022. C HD was defined per ACC/AHA guidelines as ≥50% stenosis in major epicardial arteries on angiography or prior myocardial infarction/revascularization.

Exclusion criteria: (1) severe infection; (2) active malignancy (pathologically confirmed); (3) severe liver failure (Child-Pugh C); (4) severe renal failure (eGFR <30 mL/min/1.73m²).Patient selection is shown in **Figure 1**.

**Figure 1.**
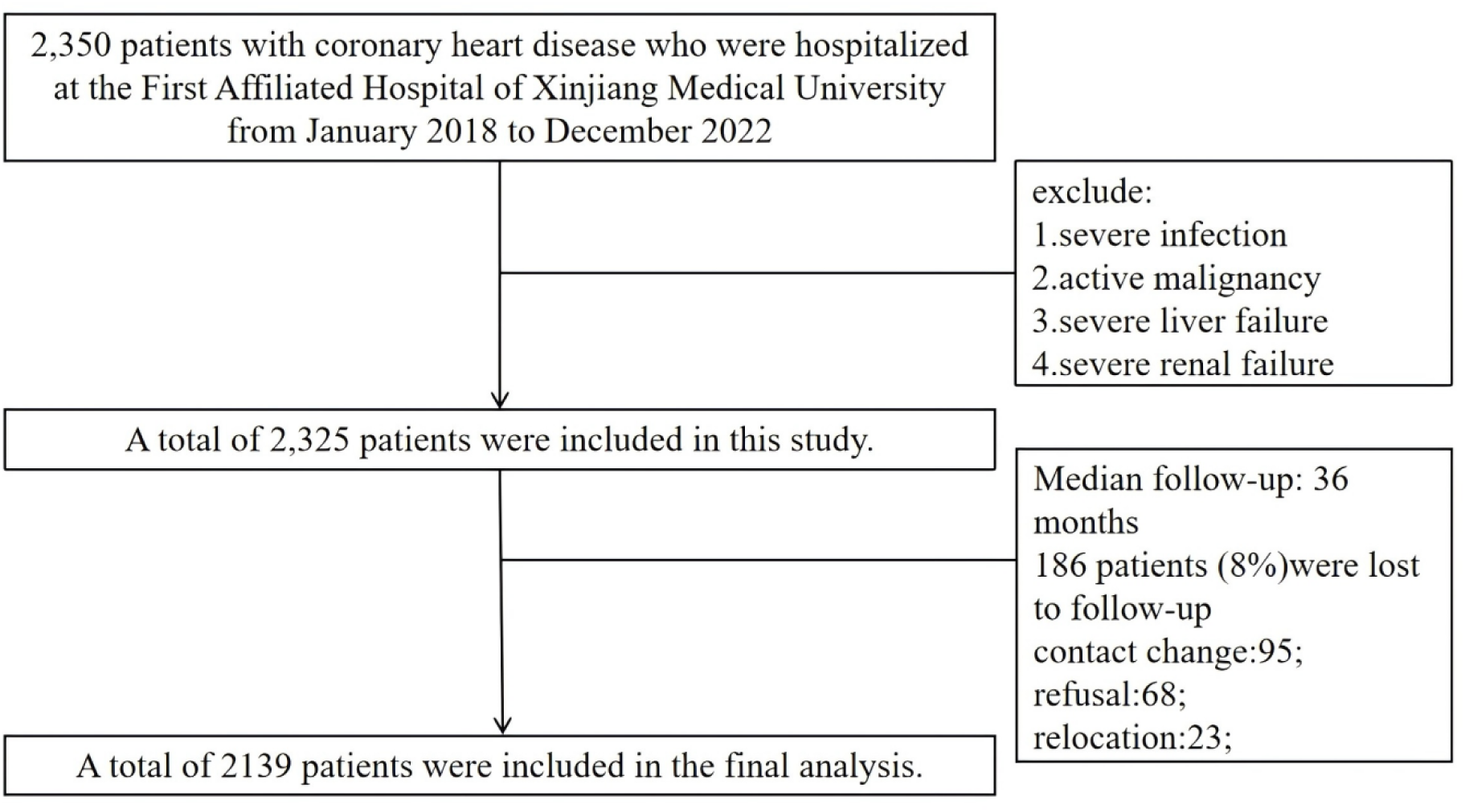
flow chart.

The study was approved by the institutional ethics committee (K202508-19) a nd conducted per the Declaration of Helsinki. Informed consent was waived given the retrospective design.

### Data collection

Baseline data were extracted from electronic medical records, including demographics, comorbidities, laboratory measurements, echocardiography, functional assessments, and discharge medications.

#### Sociodemographic characteristics

Age, sex, BMI, smoking history.

#### Comorbidities were defined per established criteria

Diabetes: fasting glucose ≥7.0 mmol/L, OGTT ≥11.1 mmol/L, HbA1c ≥6.5%, or glucose-lowering medications (ADA criteria)(8);

Hypertension: BP ≥140/90 mmHg or antihypertensive use(9);

Heart failure: clinical syndrome with elevated natriuretic peptides (NT-proBNP> 125 pg/mL or BNP >35 pg/mL) and/or structural/functional abnormality onech o (universal definition). LVEF by Simpson’s biplane method: HFrEF ≤40%,HF mrEF 41-49%, HFpEF ≥50%(10);

CKD: eGFR <60 mL/min/1.73m² (CKD-EPI equation) or ACR >30 mg/g (KDIGO)(11);

Anemia: Hb <130 g/L (men) or <120 g/L (women)(12);

Atrial fibrillation: documented on ECG, Holter, or history; Functional assessments by trained staff:

Cognitive: MoCA <26(13);

Motor: Fugl-Meyer <66(14);

Hearing: pure-tone audiometry >25 dB HL(15);

Visual: Snellen acuity <0.5(16);

Frailty: FRAIL scale (0=robust, 1-2=pre-frail, ≥3=frail)(17);

ADL: Katz index ≤2=dependence(18);

#### Laboratory

Hemoglobin, platelets, albumin, lipids, creatinine (eGFR), NT-pro BNP from fasting samples within 24 hours of admission.

#### Medications at discharge

Antiplatelets, anticoagulants, statins, beta-blockers, RAS inhibitors, diuretics.

#### Follow-up and outcomes

Trained staff conducted telephone interviews every 3 months, collecting fall d etails (date, location, circumstances, injury, medical attention). Follow-up: discharge to December 31, 2023 (median 36 months, IQR 24-48).

Of 2,325 enrolled, 186 (8.0%) were lost to follow-up (contact change 51.1%, refusal 36.6%, relocation 12.4%). Baseline characteristics were similar between completers and lost patients, indicating non-differential attrition. Final analysis: 2,139 patients (92.0% completion).

Clinical events were adjudicated by two blinded cardiologists using medical records, discharge summaries, death certificates, and interviews. A third cardiologist resolved disagreements.

#### Primary outcomes

All-cause mortality(ACM)(19); cardiac death(CM)(20).

#### Secondary outcomes

MACE: cardiac death, non-fatal MI, target vessel revascularization(21).

MACCE: MACE and non-fatal stroke(22).

### Statistical analysis

Analyses used R 4.4.1; *P*<0.05 significant. Continuous variables: mean±SD or median (IQR); categorical: frequencies (%). Comparisons by t-test/Mann-Whit ney U and chi-square/Fisher’s exact test. Kaplan-Meier curves with log-rank te st compared survival.

Cox regression: Variables with univariate *P*<0.05 plus age/sex entered multivariable models: Model 1 (age, sex); Model 2 (+comorbidities); Model 3 (+func tional status); Model 4 (+LVEF, eGFR, hemoglobin, diuretics). Proportional hazards tested via Schoenfeld residuals; violations handled by stratified models. Subgroups: Age, sex, HF, diabetes, frailty, LVEF, CKD, cognition; interaction s via multiplicative terms.

Sensitivity analyses: (1) Fine-Gray competing risk models; (2) landmark analy sis (>6 months); (3) IPTW with propensity score (14 covariates), stabilized weights (1st-99th percentile truncation), SMD<0.10; (4) excluding LVEF<40%, eGFR<30, age≥85.

## Results

### Baseline characteristics

A total of 2,139 patients with coronary heart disease were included in the final analysis, of whom 171 (8.0%) experienced at least one fall during follow-u p. Baseline characteristics stratified by fall status are presented in **Table 1**.

**Table 1.**
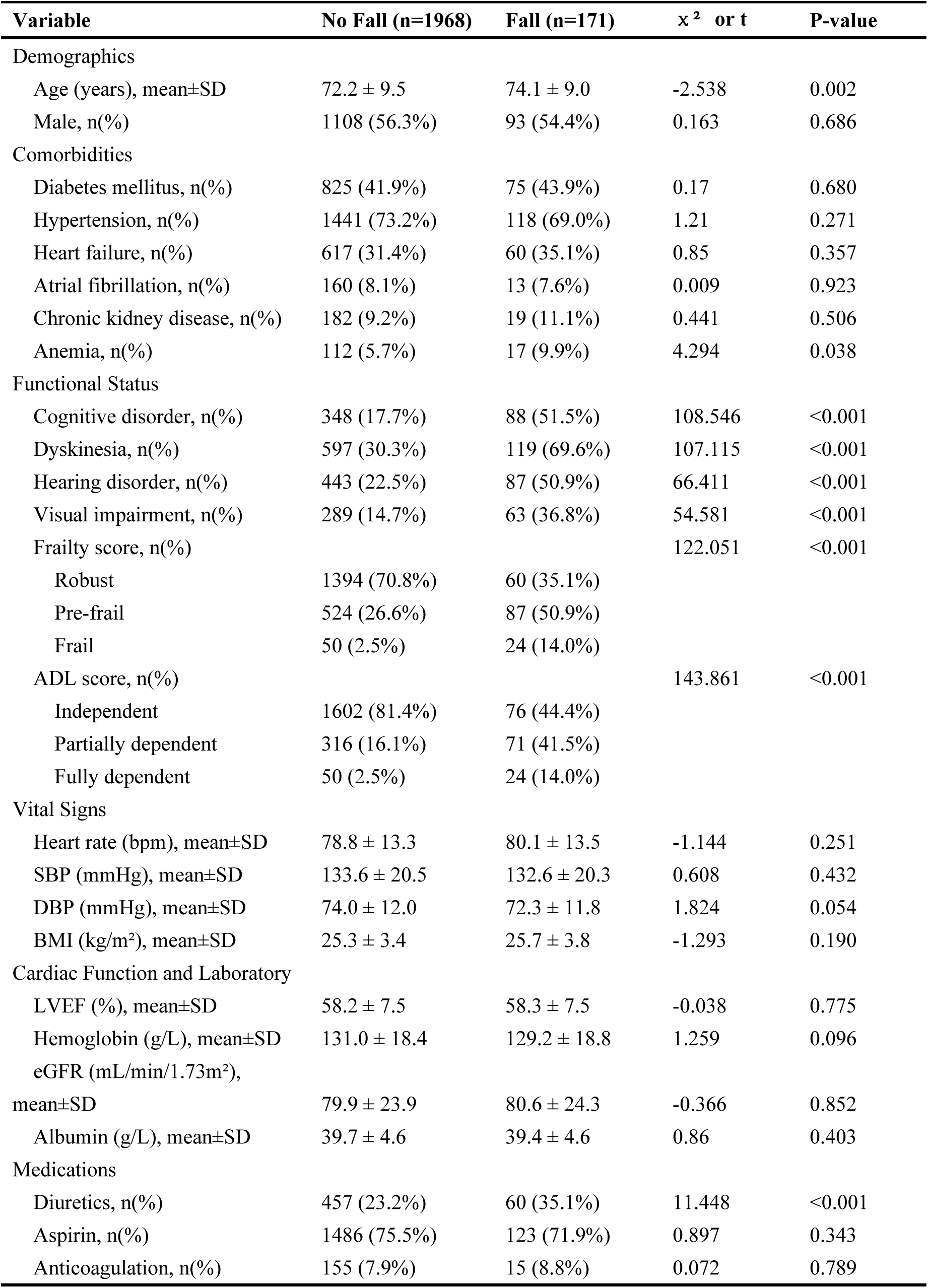
Baseline characteristics.

Patients who experienced falls were significantly older (74.1±9.0 vs. 72.2±9.5 years, *P*=0.002) and had higher prevalence of anemia (9.9% vs. 5.7%, *P*=0.03 8) and diuretic use (35.1% vs. 23.2%, *P*<0.001). Marked differences were observed in functional status: compared with non-fallers, patients with falls had significantly higher rates of cognitive impairment (51.5% vs. 17.7%), dyskinesia (69.6% vs. 30.3%), hearing disorder (50.9% vs. 22.5%), visual impairment ( 36.8% vs. 14.7%), and ADL dependence (all *P*<0.001). Frailty distribution als o differed significantly between groups (*P*<0.001): only 35.1% of fallers were robust, compared with 70.8% of non-fallers. Other baseline characteristics, including sex, BMI, comorbidities (diabetes, hypertension, heart failure, atrial fibrillation, CKD), and cardiac function (LVEF) were similar between groups.

### Survival analysis

Kaplan-Meier survival curves (**Figure 2**) demonstrated significantly lower cumulative event-free survival in patients with falls compared with those without falls for all endpoints. The log-rank test confirmed significant differences for all-cause mortality (*P*<0.001), cardiac mortality (*P*<0.001), MACE (*P*<0.001), and MACCE (*P*<0.001).

**Figure 2.**
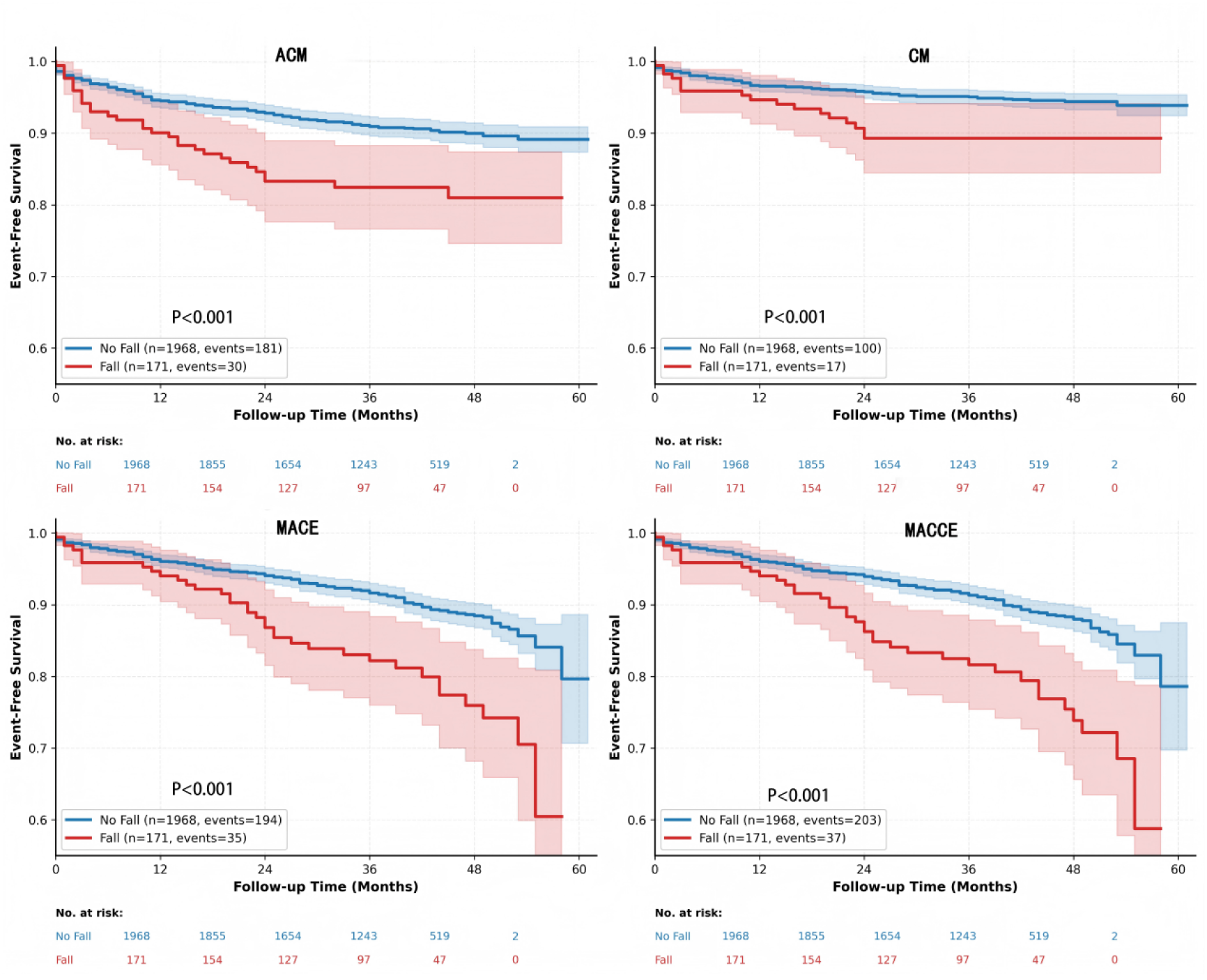
Kaplan-Meier Survival Curves for Clinical Outcomes Stratified by Fall Status.

**Figure 3.**
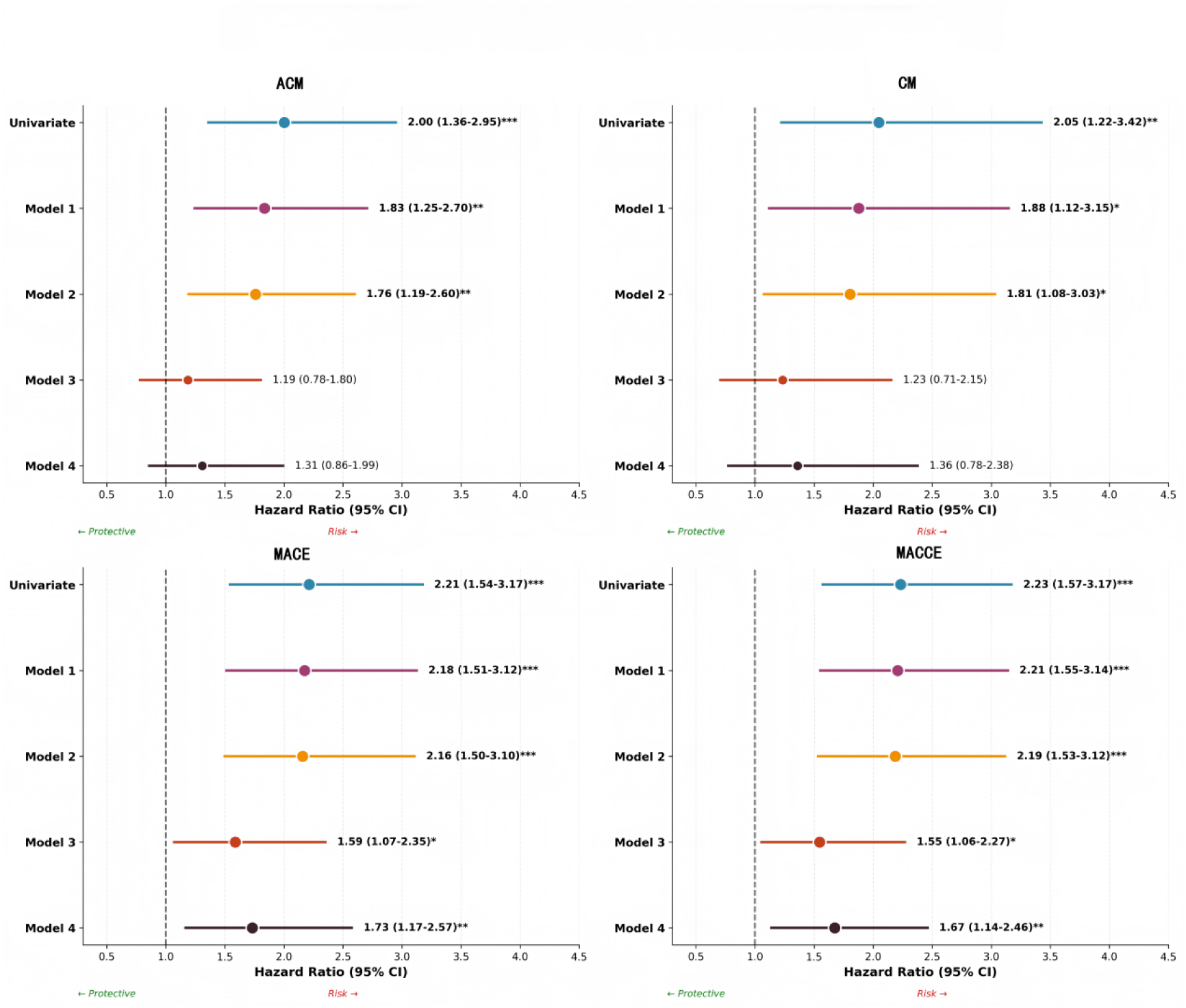
Multivariate Cox regression forest plot.

### Univariate Cox regression analysis

In univariate analysis (**Supplementary Table 1**), history of falls was significantly associated with increased risk across all endpoints. Patients who experienced falls had approximately twice the risk of all-cause mortality (*HR*=2.00, 95% *CI*: 1.36–2.95, *P*<0.001), cardiac mortality (*HR*=2.05, 95%*CI*: 1.22–3.42, *P*=0. 006), MACE (*HR*=2.21, 95%*CI*: 1.54–3.17, *P*<0.001), and MACCE (*HR*=2.23, 95%*CI*: 1.57–3.17, *P*<0.001).

Additional significant prognostic factors included functional status indicators (dyskinesia, frailty score, ADL score, cognitive impairment, and hearing disorder), traditional cardiovascular risk factors (heart failure, CKD, LVEF, NT-proBNP), and diuretic use (all *P*<0.05).

### Multivariable Cox regression analysis

Multivariable Cox regression with stepwise adjustment revealed differential associations between falls and various outcomes (**Table 2**). After adjusting for functional status (Model 3), associations with mortality were substantially attenuated and became non-significant (all-cause mortality: *HR*=1.19, *P*=0.422; cardiac mortality: *HR*=1.23, *P*=0.459), whereas associations with cardiovascular eve nts remained robust (MACE: *HR*=1.59, *P*=0.021; MACCE: *HR*=1.55, *P*=0.025)

**Table 2.** Association Between Falls and Clinical Outcomes: Multivariable Cox Regression Analysis with Stepwise Adjustment.

| Model | ACM HR<br>(95% CI) | ACM<br>P-value | CM HR<br>(95% CI) | CM<br>P-value | MACE HR<br>(95% CI) | MACE<br>P-value | MACCE HR<br>(95% CI) | MACCE<br>P-value |
| --- | --- | --- | --- | --- | --- | --- | --- | --- |
| Unadjusted | 2.00 (1.36-2.95) | <0.001 | 2.05 (1.22-3.42) | 0.006 | 2.21 (1.54-3.17) | <0.001 | 2.23 (1.57-3.17) | <0.001 |
| Model 1 | 1.83 (1.25-2.70) | 0.002 | 1.88 (1.12-3.15) | 0.017 | 2.18 (1.51-3.12) | <0.001 | 2.21 (1.55-3.14) | <0.001 |
| Model 2 | 1.76 (1.19-2.60) | 0.004 | 1.81 (1.08-3.03) | 0.025 | 2.16 (1.50-3.10) | <0.001 | 2.19 (1.53-3.12) | <0.001 |
| Model 3 | 1.19 (0.78-1.80) | 0.422 | 1.23 (0.71-2.15) | 0.459 | 1.59 (1.07-2.35) | 0.021 | 1.55 (1.06-2.27) | 0.025 |
| Model 4 | 1.31 (0.86-1.99) | 0.210 | 1.36 (0.78-2.38) | 0.281 | 1.73 (1.17-2.57) | 0.006 | 1.67 (1.14-2.46) | 0.009 |

Model 1: Adjust age and gender;
Model 2: Model 1 + Comorbidities (Diabetes, Hypertension, Heart failure, Atrial fibrillation, CKD, anemia);
Model 3: Model 2 + Function Status (Cognitive Impairment, Motor Impairment, Weakness Score);
Model 4: Full adjustment model (including LVEF, eGFR, hemoglobin, and diuretics).

In the fully adjusted model (Model 4), falls independently predicted MACE (*HR*=1.73, 95%*CI*: 1.17–2.57, *P*=0.006) and MACCE (*HR*=1.67, 95%*CI*: 1.14–2.46, *P*=0.009), but not mortality outcomes (ACM: *HR*=1.31, *P*=0.210; CM: *HR*=1.36, *P*=0.281).

### Subgroup analysis

Subgroup analyses are shown in **Table 3** and **Supplementary Figure1,2.** The associations between falls and MACE/MACCE were consistent across age, se x, heart failure status, cardiac function, and cognitive impairment subgroups (a ll *P* for interaction >0.05). However, frailty status significantly modified these associations (*P* for interaction <0.001): among robust patients, falls were associated with markedly elevated risks (MACE: *HR*=4.08, 95%*CI*: 2.37–7.01; MACCE: *HR*=3.94, 95%*CI*: 2.30–6.76), whereas among pre-frail/frail patients, associations were non-significant (MACE: *HR*=1.15, *P*=0.576; MACCE: *HR*=1.1 9, *P*=0.470).

**Table 3.** Subgroup analyses of MACE and MACCE using Cox regression.

| Subgroup | Category | MACE<br>(N) | MACE<br>(Events) | MACE HR (95%<br>CI) | P-value | P interaction | MACCE<br>(N) | MACCE<br>Events | MACCE HR (95%<br>CI) | P-value | P interaction |
| --- | --- | --- | --- | --- | --- | --- | --- | --- | --- | --- | --- |
| Age | <75 years | 1178 | 115 | 2.02 (1.16-3.54) | 0.014 | 0.781 | 1178 | 123 | 2.16 (1.28-3.66) | 0.004 | 0.988 |
|  | ≥75 years | 961 | 114 | 2.24 (1.40-3.60) | <0.001 |  | 961 | 117 | 2.17 (1.36-3.48) | 0.001 |  |
| Gebder | Male | 1201 | 124 | 2.55 (1.56-4.16) | <0.001 | 0.437 | 1201 | 130 | 2.57 (1.59-4.14) | <0.001 | 0.427 |
|  | Female | 938 | 105 | 1.91 (1.12-3.26) | 0.017 |  | 938 | 110 | 1.93 (1.15-3.24) | 0.013 |  |
| Heart failure | No | 1462 | 121 | 2.12 (1.27-3.55) | 0.004 | 0.985 | 1462 | 129 | 2.24 (1.38-3.65) | 0.001 | 0.833 |
|  | Yes | 677 | 108 | 2.14 (1.29-3.55) | 0.003 |  | 677 | 111 | 2.08 (1.25-3.44) | 0.005 |  |
| Diabetes<br>mellitus | No | 1239 | 130 | 2.64 (1.67-4.18) | <0.001 | 0.250 | 1239 | 137 | 2.75 (1.77-4.27) | <0.001 | 0.162 |
|  | Yes | 900 | 99 | 1.71 (0.95-3.07) | 0.072 |  | 900 | 103 | 1.63 (0.91-2.92) | 0.099 |  |
| Frailty status | Robust | 1454 | 123 | 4.08 (2.37-7.01) | <0.001 | <0.001 | 1454 | 127 | 3.94 (2.30-6.76) | <0.001 | <0.001 |
|  | Pre-frail/Frail | 685 | 106 | 1.15 (0.71-1.87) | 0.576 |  | 685 | 113 | 1.19 (0.74-1.89) | 0.470 |  |
| LV function | ≥50% | 1797 | 173 | 2.23 (1.48-3.37) | <0.001 | 0.885 | 1797 | 182 | 2.29 (1.54-3.40) | <0.001 | 0.802 |
|  | <50% | 342 | 56 | 2.10 (0.99-4.45) | 0.053 |  | 342 | 58 | 2.05 (0.97-4.35) | 0.060 |  |
| CKD | No | 1938 | 196 | 2.04 (1.36-3.04) | <0.001 | 0.198 | 1938 | 206 | 2.08 (1.41-3.06) | <0.001 | 0.244 |
|  | Yes | 201 | 33 | 3.79 (1.61-8.95) | 0.002 |  | 201 | 34 | 3.62 (1.54-8.51) | 0.003 |  |
| cognitive<br>disorder | No | 1703 | 168 | 2.03 (1.18-3.51) | 0.011 | 0.957 | 1703 | 172 | 1.98 (1.15-3.43) | 0.014 | 0.927 |
|  | Yes | 436 | 61 | 1.99 (1.17-3.38) | 0.011 |  | 436 | 68 | 1.92 (1.16-3.17) | 0.011 |  |

Notable findings in other subgroups included higher fall-associated MACE ris k in non-diabetic versus diabetic patients (HR=2.64 vs. 1.71, P for interaction =0.250) and particularly elevated risk among CKD patients (*HR*=3.79, 95%*CI*: 1.61–8.95, *P*=0.002).

### Sensitivity analyses

Multiple sensitivity analyses confirmed the robustness of primary findings (**Supplementary Table2** and **Supplementary Figure3**). Results remained consistent across competing risk models (MACE: *HR*=1.77, *P*=0.005), landmark analysis excluding early events (*HR*=1.84, *P*=0.007), IPTW analysis (*HR*=2.09, *P*=0.022), a nd sequential exclusion of patients with LVEF<40% (*HR*=1.64, *P*=0.018), eGF R<30 (*HR*=1.66, *P*=0.015), or age≥85 years (*HR*=1.87, *P*=0.002).

## Discussion

The present study reveals distinct pathways through which falls influence different outcomes in patients with coronary heart disease. After adjustment for functional status, the associations between falls and both all-cause and cardiac mortality were attenuated and no longer significant, indicating that functional status—particularly frailty and dyskinesia—mediates the relationship between falls and mortality. In contrast, falls remained independently associated with MACE (*HR*=1.73, 95%*CI*: 1.17–2.57, *P*=0.006) and MACCE (*HR*=1.67, 95%*CI*: 1.14–2.46, P=0.009) even after full adjustment, suggesting direct triggering of cardiovascular events. Subgroup analysis demonstrated that frailty status significantly modified this association—falls conferred fourfold increased MACE/MACCE risk in robust patients but not in frail patients. These findings offer a novel evidence-based perspective for individualized risk management.

Univariate analysis revealed significant associations between falls and both all-cause and cardiac mortality (*HR* ≈ 2.0), which were fully attenuated after adjusting for functional status indicators. This indicates that falls increase mortality risk indirectly through exacerbating functional decline. Previous studies support this interpretation, demonstrating that falls lead to ADL decline and accelerate frailty progression, both established predictors of mortality(23–25). A large-scale study confirmed that complications following geriatric trauma, rather than trauma itself, independently predicted mortality(26). In our analysis, when frailty and ADL impairment were included in the model, the direct effect of falls on mortality was substantially diminished. These findings explain inconsistent conclusions in previous studies that neglected functional status as a key mediator.

Unlike mortality, falls remained independently associated with MACE and MACCE in fully adjusted models, with consistent results across seven sensitivity analyses (*HR* range: 1.64–2.09). This effectively excluded reverse causation, competing risks, or results driven by specific subgroups. The persistently elevated hazard ratios after full adjustment serve as warning signals for subsequent cardiovascular events, distinguishing the fall-cardiovascular event pathway from the fall-mortality pathway. Mechanistically, acute stress can injure the myocardium through multiple pathways: increased cortisol and catecholamines activate the autonomic nervous system, triggering inflammatory responses and DAMP activation(27). Additionally, trauma induces a hypercoagulable state with increased thrombotic risk(28). In the cerebrovascular domain, acute stress increases stroke risk through adrenaline-mediated platelet activation, Factor XII-mediated coagulation cascade initiation, and TLR4-mediated neuroinflammation(29). These convergent mechanisms explain why falls were similarly associated with MACCE, which encompasses stroke. Frailty status significantly modified the fall-cardiovascular event association (*P* for interaction <0.001). Among robust patients, falls conferred markedly elevated risk (MACE: *HR*=4.08, 95%*CI*: 2.37–7.01; MACCE: *HR*=3.94, 95%*CI*: 2.30–6.76), whereas among pre-frail/frail patients, associations were non-significant (MACE: *HR*=1.15, 95%*CI*: 0.71–1.87; MACCE: *HR*=1.19, 95%*CI*: 0.74–1.89).

This paradoxical finding warrants careful interpretation. First, competing risk offers an important explanation—frail patients are more susceptible to non-cardiovascular events (fracture, infection, disability) that may preclude MACE occurrence. In our study, patients with falls had high prevalence of ADL dependence (55.5%) and pre-frailty/frailty (64.9%), exemplifying this high-risk population. Second, a floor effect may be at play—frail patients already have substantially elevated baseline cardiovascular risk, limiting the detectable incremental impact of falls.

This phenomenon is not isolated to our study—previous research reported similar frailty-related effect modification, such as elevated blood pressure paradoxically protecting against mortality in frail elderly(30). These findings carry important clinical implications. Falls in robust coronary heart disease patients should prompt comprehensive cardiovascular evaluation (electrocardiography, cardiac enzymes, ambulatory ECG monitoring) given the fourfold MACE/MACCE risk elevation. For frail patients, clinicians should prioritize both cardiovascular assessment and vigilance for non-cardiovascular complications, while recognizing that conventional statistical frameworks may underestimate cardiovascular consequences in this population.

Other subgroup findings. In the diabetes subgroup, MACE risk elevation following falls was higher in non-diabetic patients (*HR*=2.12) than diabetic patients (*HR*=1.31), though interaction was non-significant (*P*=0.089). This may reflect blunted stress responses from diabetic autonomic neuropathy or a ceiling effect. Among CKD patients, falls conferred markedly increased MACE risk (*HR*=3.79, 95%*CI*: 1.61– 8.95)—their hypercoagulable state and severe vascular calcification likely render them particularly vulnerable to stress-triggered thrombotic events. Other subgroups (age, sex, heart failure, cardiac function, cognitive function) showed consistent associations, reinforcing robustness of primary findings.

Clinical recommendations. Our findings support several recommendations. First, falls should be incorporated into comprehensive risk assessment frameworks; clinicians should routinely inquire about recent falls during follow-up. Second, robust patients experiencing falls require comprehensive cardiovascular evaluation with consideration of intensified secondary prevention (optimizing antiplatelet therapy, lipid management, blood pressure control). Third, frail patients need attention to both cardiovascular and non-cardiovascular complications post-fall, including fracture risk evaluation, infection prevention, and rehabilitation. Fourth, future interventional studies should stratify by frailty status to clarify differential benefits of fall prevention strategies, aligned with World Guidelines for Falls Prevention(31).

This single-center retrospective study has limited generalizability and cannot establish causality despite adjusting for multiple confounders. The non-significant fall-MACE association in frail patients may reflect limited sample size and competing risks; future studies should employ competing risk models for validation. Falls were ascertained via telephone follow-up, susceptible to recall and underreporting bias. Mechanistic discussions remain speculative without direct biomarker measurements.

## Conclusion

Falls independently predict long-term MACE and MACCE in coronary heart disease patients, with mortality effects mediated by functional status. Frailty significantly modifies this association—robust patients experiencing falls face substantially elevated cardiovascular risk and warrant comprehensive evaluation. These findings provide evidence-based support for individualized fall management and prognostic intervention in coronary heart disease.

## Author contributions

Changjiang Deng: Conceptualization, Data curation, Formal analysis, Methodology, Software, Visualization, Writing-original draft. Yuhang Men: Conceptualization, Data curation, Formal analysis, Methodology, Validation, Writing-original draft. Xinshu Xu: Investigation, Validation. Yujie Pang: Investigation, Visualization. Zihan Tang: Investigation, Visualization. Haowen Ren: Investigation, Visualization. Weiyan Cui: Resources, Supervision, Writing-review & editing. Junrui Hou: Data curation, Investigation. Maimaitiming·Muyesaier: Data curation, Investigation. Zhihang Chen: Formal analysis, Software. Haoyu Chen: Investigation, Validation. Ting-Ting Wu: Conceptualization, Methodology, Project administration, Resources, Supervision, Writing-review & editing.

## Data availability statement

Data are available from the corresponding author upon reasonable request.

## Supplementary

Supplementary Table1: Univariate Cox Regression Analysis of Risk Factors for Clinical Outcomes.

Supplementary Table2: Sensitivity Analyses: Association Between Falls and Cardiovascular Events.

Supplementary Figure1: Subgroup Analysis: Association Between Falls and MACE. Supplementary Figure2: Subgroup Analysis: Association Between Falls and MACCE. Supplementary Figure3: Sensitivity analysis forest plot.

